# Student-run Clinic Participation and Likelihood of Practicing Primary Care: A Meta-Analysis

**DOI:** 10.64898/2026.01.24.26344631

**Authors:** Nicholas Peoples, Shangzhi Xiong, Simone Nguyen, Daniel C. Brock, Dana Clark

## Abstract

**Importance:** The United States is facing a projected shortage of 40,000 primary care physicians by 2034. Student-run clinics (SRCs) are widely regarded as service-learning environments that may encourage students to enter a primary care specialty, but prior studies have yielded conflicting results and are limited to non-generalizable, single site analyses.

**Objective:** To compare likelihood of practice in a primary care specialty between students who did and did not participate in an SRC via pooled meta-analysis of U.S.-based studies.

**Study Selection:** Studies were first identified through a comprehensive library of U.S.-based SRC literature. The inclusion criteria were publications on SRCs in the United States with MD/DO students, from all time until March 1, 2024. Exclusion criteria were: full text not available; published abstract/textbook/dissertation/thesis; not in English. Two authors independently screened the database for publications on SRC participation and practice in primary care specialties. To identify relevant literature after March 1, 2024, the authors performed iterative snowball sampling of the bibliographies of included studies and their Google Scholar “cited by” lists until saturation. Finally, we included original data from the single largest study on this topic.

**Data Extraction and Synthesis:** We evaluated study quality using the NIH Study Quality Appraisal Tool. We used a random-effects model to account for heterogeneity.

**Main Outcomes and Measures:** The primary outcome was the relative likelihood (risk ratio) of pursuing a primary care specialty among SRC volunteers versus non-volunteers. We used a funnel plot and sensitivity analysis to assess for bias.

**Results:** Seven studies met inclusion criteria with a cumulative sample size of 7,468 students. SRC volunteers pursued primary care at 102% to 160% the rate of non-volunteers. The pooled risk ratio was 1.25 (95% CI: 1.09–1.44). Funnel plot and multiple sensitivity analyses did not suggest publication bias or undue influence from included studies.

**Conclusions and Relevance:** SRC participation is associated with a statistically-significant 25% increased likelihood of practicing in a primary care specialty. These findings may inform national and institutional strategies to support service-learning and address the national primary care workforce shortage.

**KEY POINTS:** *Question:* Is participation in a student-run clinic during medical school associated with increased likelihood of practicing in a primary care specialty?

*Findings:* Despite conflicting results in the literature among small and single-site studies, in this meta-analysis of 7,468 medical students, participation in a student-run clinic was associated with a statistically significant 25% increased likelihood of practicing in a primary care specialty.

*Meaning:* Student-run clinics may be a potential strategy for strengthening the pipeline into primary care and reducing the projected shortage of 40,000 primary care physicians.

## Introduction

The Association of American Medical Colleges projects a shortage of 20,200 to 40,400 primary care (PC) physicians in the United States by 2034.^1^ Student-run clinics (SRC) have been recognized as a potential pipeline for the PC workforce,^2^ where service learning, increased autonomy, and early PC exposure might positively influence students toward a PC career.^2-5^ Others caution, however, that moral injury in the setting of limited resources can dissuade students from this path.^3^

Empiric work provides mixed results.^2,4-15^ The classic 1985 Campos-Outcalt study^2^ continues to be cited—even decades later—as evidence of a positive relationship. Some recent studies agree,^4-6^ but even more studies have not found an association.^7-11^ It is difficult to draw broader conclusions as each study is single-site and not generalizable.

Others have studied whether the presence of an SRC is associated with increased institutional match rates into PC.^12,13^ Unfortunately, these studies cannot capture whether students who volunteer at SRCs pursue PC more often. Some reviews also exist.^14-16^ However, they do not restrict their search to the United States. Since the incentives, opportunity costs for choosing PC, and the medical education and healthcare systems vary markedly across countries, these differences introduce potential confounding.

Finally, there is a general need of more representative data for SRCs. A 2025 scoping review comprehensively analyzed the U.S. literature on SRCs and found a preponderance of low quality, non-generalizable data from a handful of medical schools.^17^ Currently, only one SRC meta-analysis has been attempted.^18^

Therefore, we performed a meta-analysis to better assess the relationship of SRC participation and likelihood of practicing PC. We further include original data from the largest individual study on this topic in our pooled analysis.^19^

## Methods

### Article Selection

We used the comprehensive “library” of SRC literature from Peoples et al. 2025 in *Academic Medicine*.^17^ This review was inclusive of all publications on SRCs in the United States with MD/DO students from all time until March 1, 2024. Detailed results are available within the source publication.^17^

Two authors, AA and BB, independently screened the library (n=503 publications) by title/abstract and full text. They achieved a percent agreement of 99.6%, with AA identifying 14 papers and BB independently identifying 12 of the same articles (agreement on 501/503 items). They performed three rounds of snowball sampling in July 2025, iteratively reviewing the bibliographies and “cited by” lists for each paper on Google Scholar until no new relevant publications could be identified. They achieved a percent agreement of 100% for each round.

### Data Extraction

AA and BB extracted data from each included publication, such as study design, sample size, and sociodemographic data. We used the NIH Study Quality Appraisal Tool^20^ to assess the methodologic quality of each study.

### Meta-Analysis

We dichotomized “exposure” (volunteering at an SRC vs. not) and “outcome” (practicing PC vs. not) from included studies, pooling the results for the risk ratio of practicing PC between SRC volunteers and non-volunteers. We defined “likelihood of PC practice” to be inclusive of three different timepoints in the PC career pipeline: declared career choice, match results, and PC practice years after residency. We used a random effects model because the study settings and design are heterogenous. We conducted multiple sensitivity analysis and constructed a funnel plot (Egger’s test deferred given n<10 studies) to assess for robustness and publication bias.

## Results

From 503 publications, we initially identified six viable for meta-analysis.^2.4,5,7-9^ We then included original data from Brock et al. 2026^19^ for a total of seven included studies. One measured students’ declared career choice of PC.^8^ Five measured PC match rates.^2,4,7,9,19^ One measured MDs practicing in a PC specialty 2-5 years after residency.^5^ The full search process is described in **Figure 1**. Selected characteristics of included studies are described in **Table 1**.

**Table 1.** Selected Study Characteristics. *Original, unpublished data contributed for this study, available as a pre-print.^19^ **Relationship between SRC participation and primary care identified in study. ***Quality Rating via NIH Quality Appraisal Tool; PC = Primary Care; SRC = Student-run Clinic.

| Reference | Study Method | Study Period | Sample Size | Response Rate | Outcome Type | Mean Age | Sex (%F:M) | Entered PC: SRC Volunteers (n, %) | Entered PC: Non-volunteers (n, %) | Study Result** | Quality Rating*** |
| --- | --- | --- | --- | --- | --- | --- | --- | --- | --- | --- | --- |
| **Campos-Outcalt 1985** | Retrospective | 1978-1982 | 480 | N/A | Match Results | N/A | N/A | 28 (28/29, 96.5%) | 273 (273/451, 60.5%) | Positive | Low |
| **Vaikunth et al 2014** | Retrospective | Fall 2005-Spring 2012 | 689 | N/A | Match Results | N/A | N/A | 73 (73/141, 52%) | 278 (278/548, 51%) | None | Moderate |
| **Weinreich et al 2015** | Prospective Cross-sectional Survey | June 4, 2012 to March 1, 2013 | 425 | 20.70% | Match Results | N/A | 61.7:38.3 | 121 (121/241, 50.21%) | 89 (89/184, 48.46%) | None | Moderate |
| **Brown et al 2017** | Cross-sectional Survey | November 2014 - December 2014 | 135 | 39.80% | Declared Choice of PC Career | 24.56 | 50.4:49.6 | 46 (46/78, 58.97%) | 25 (25/56, 44.64%) | None | Moderate |
| **Shah et al 2018** | Retrospective | 2004 - 2008 | 410 | N/A | PC Practice 2-5 Years After Residency | N/A | 52.5:46.4 | 84 (84/276, 30.43%) | 29 (29/134, 21.64%) | Positive | Low |
| **Thomson et al 2022** | Retrospective | 2015 - 2021 | 506 | N/A | Match Results | N/A | N/A | 120 (120/254, 47.2%) | 92 (92/252, 36.5%) | Positive | Moderate |
| **Brock et al 2026 \*** | Retrospective | 2014 - 2025 | 4,823 | N/A | Match Results | N/A | N/A | 477 (477/1157, 41.23%) | 1234 (1234/3666, 33.66%) | Positive | High |

**Figure 1.**
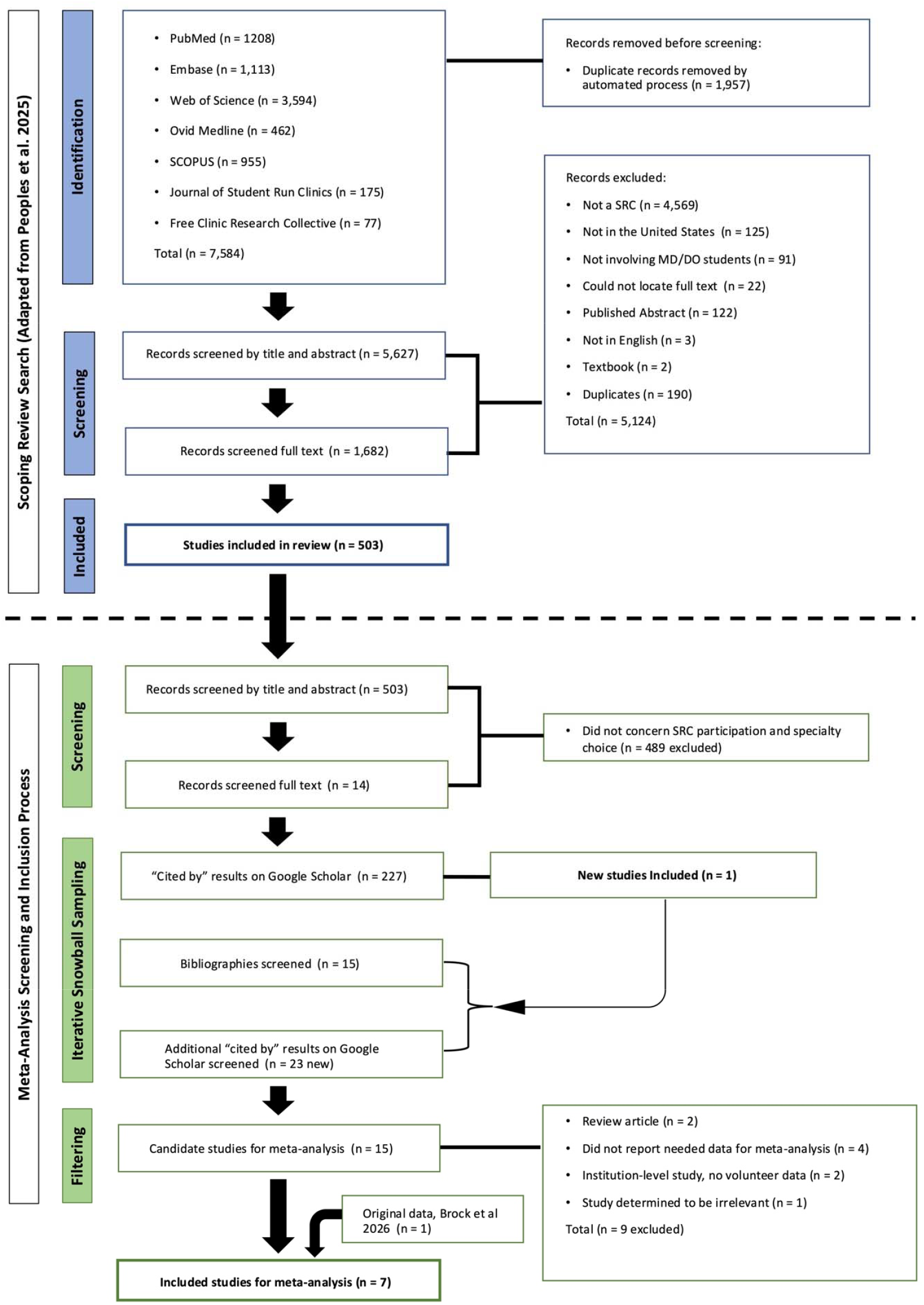
Article Search and Inclusion/Exclusion Process.

The cumulative sample size was 7,468 students. SRC volunteers pursued PC at 102%—160% the rate of non-volunteers, with a statistically-significant mean of 125% [95% CI: 1.09 – 1.44] (**Figure 2**). Heterogeneity was high (I^2^ = 79.9%). The funnel plot did not suggest significant publication bias (**Supplemental Content 1**). Results remained statistically significant across multiple sensitivity analyses (**Supplemental Content 2**).

**Figure 2.**
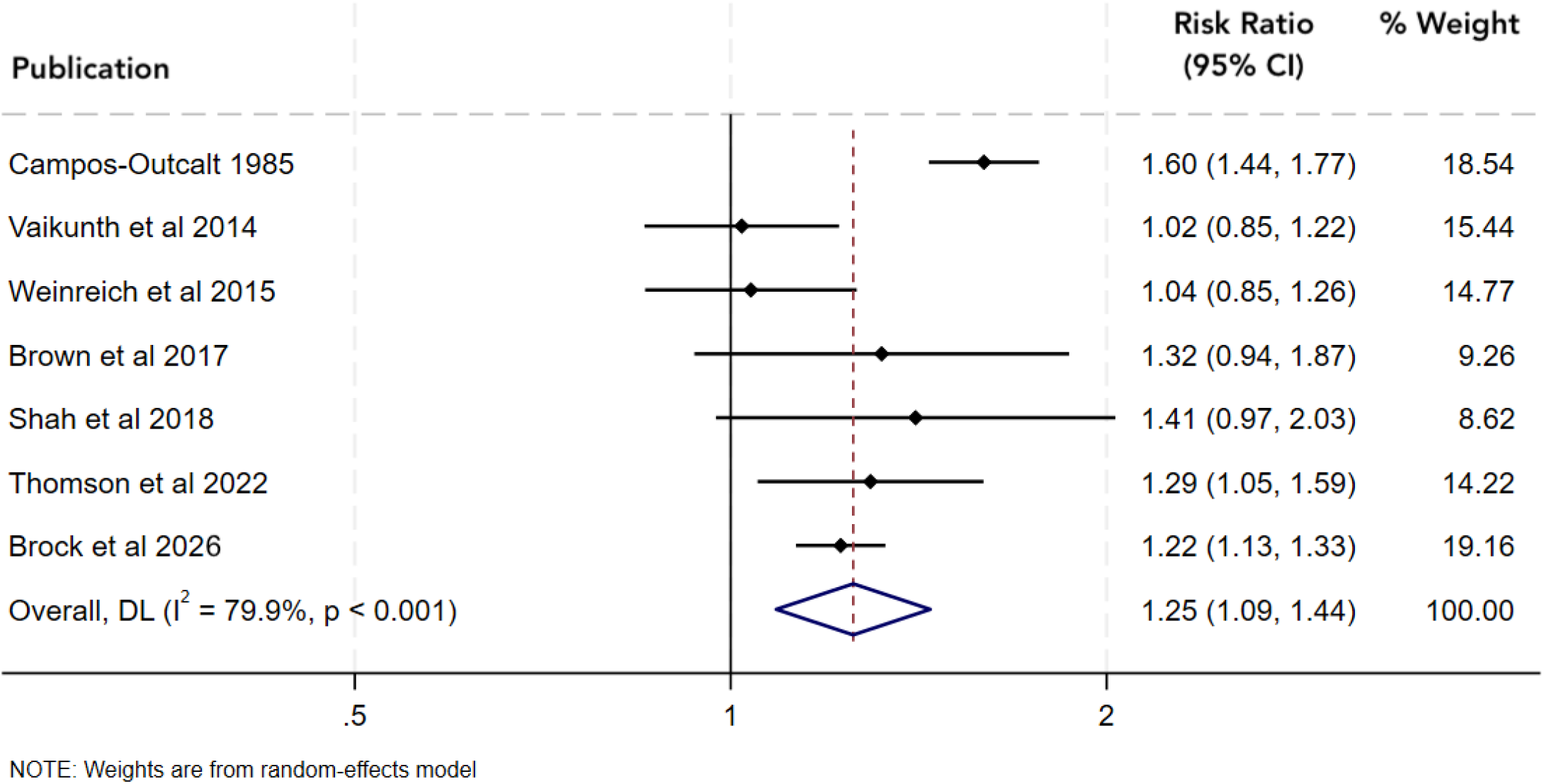
Meta-analysis of Studies on Student-run Clinic Participation and Likelihood of Primary Care Practice. CI = Confidence Internal. Weights are from random effects model.

## Discussion

This study is the second ever SRC meta-analysis and the first to assess the impact of SRC participation on likelihood of PC practice. Despite conflicting results among individual studies, in this pooled analysis of 7,468 students, participation in an SRC was associated with a statistically significant 25% increased likelihood of practicing in PC.

These results highlight the potential value of SRCs as a pipeline to help address the national PC workforce shortage. They further highlight the importance of using rigorous methods, such as meta-analysis, to study SRCs. A 2023 systematic review interpreted the mixed literature to mean that “SRC participation had little apparent impact on students’ future career directions.”^16^ Yet our results suggest they do—when examined with higher-powered, multi-site analysis.

Likewise, our results underscore the challenges of studying SRCs: there are 214 to 477 operating SRCs in 2025,^20^ yet we could only identify relevant data from 13 studies^a^, meaning the overall situation is still undetermined. Comparing volunteer records against Match results is straightforward; we encourage other SRCs to publish similar data, which would make pooled analyses more powerful in the future.

This study has notable strengths. The median sample size of individual studies was n=480, but we achieved a composite sample size of n=7,468. Our study is further made robust by including data from three different timepoints in the PC career pipeline. Finally, results remained significant across six different sensitivity analyses.

There are several important limitations. First, we cannot show causality: SRCs may influence students toward PC, or students interested in PC may simply gravitate toward SRCs. Future work should attempt longitudinal, prospective studies that account for confounding variables. Pre-post assessments of early medical student career interests, frequency and duration SRC participation, and Match results could better elucidate this relationship.

Second, the methodological quality of studies is limited, with several not reporting response rates, inclusion/exclusion criteria, or using a consistent definition of “primary care.” Many were also missing sociodemographic data such as mean age, sex, and ethnicity. To mitigate this (to the extent possible), we used a random effects model to account for the high heterogeneity among studies.

Third, the funnel plot suggests slight asymmetry. While this could indicate small-study effects or publication bias, interpretation is limited by the modest number of studies and high heterogeneity. Thus, it should be interpreted cautiously. Nonetheless, potential slight asymmetry is unlikely reflect compromising bias.

Finally, we excluded four relevant studies because they did not report needed data.^6,10,11,21^ However, three found statistically-significant positive (or conditionally-positive) associations between SRC participation and PC.^6,11,21^ Therefore, the true positive association is potentially even stronger than what our results show.

## Conclusion

Despite conflicting results from individual studies, in this meta-analysis of 7,468 medical students, participation in a student-run clinic was associated with a statistically significant 25% increased likelihood of practicing in a primary care specialty. As the best available evidence to date, these findings may be useful to leaders in primary care, medical education, and policy in considering whether and how these unique clinics might be leveraged to help address the national primary care workforce shortage.

## Data Availability

The authors will gladly share the data underpinning this study upon reasonable request.

## Ethics

This is secondary research of published literature and did not require the approval of an institutional review board.

## Data Sharing Statement

The authors will gladly share the data underpinning this study upon reasonable request.

## Supplemental Content 1 – Funnel Plot

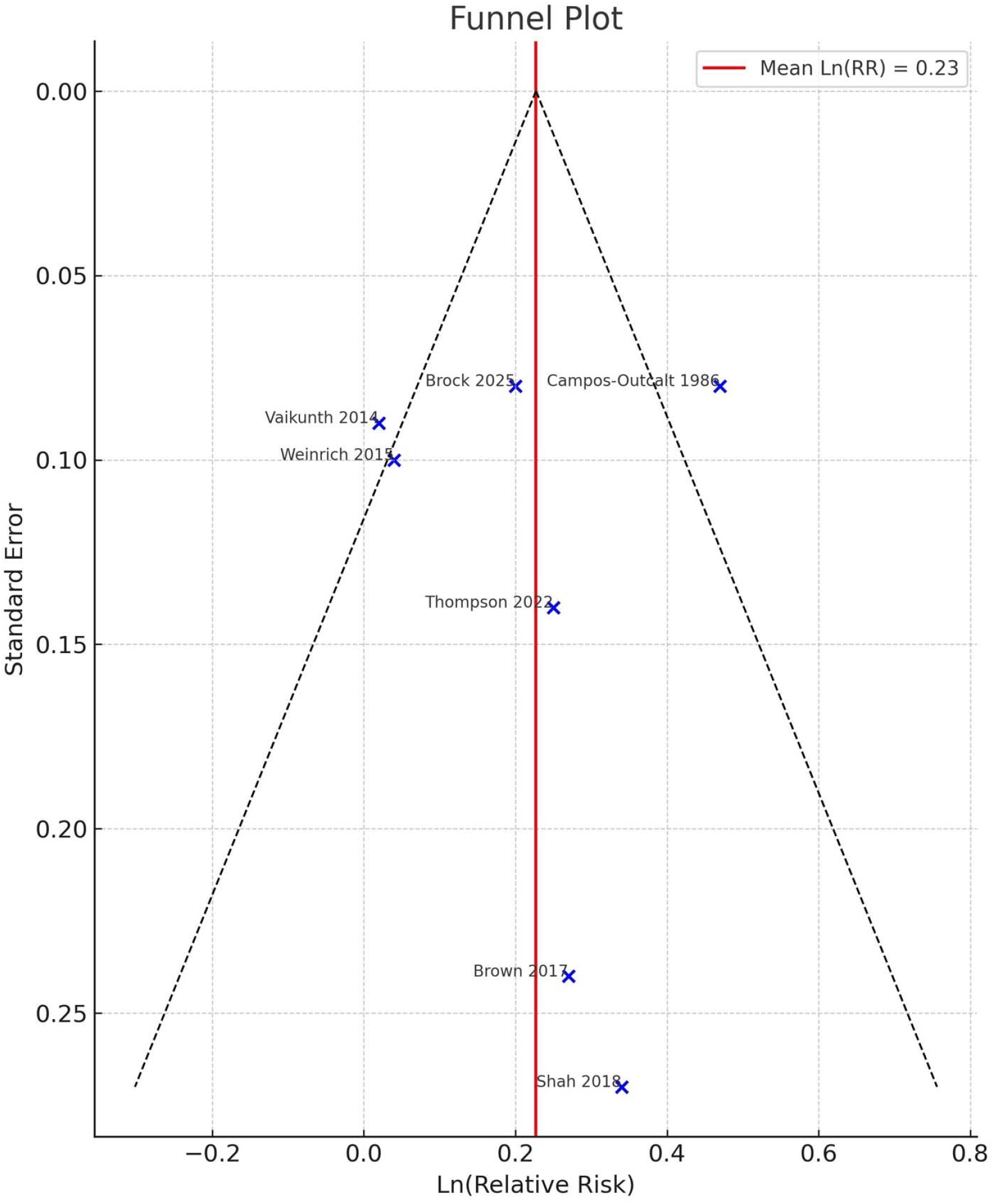

## Supplemental Content 2 – Sensitivity Analyses

**Sensitivity Analysis 1:** Brown 2017, Shah 2018, Campos-Outcalt 1985 removed

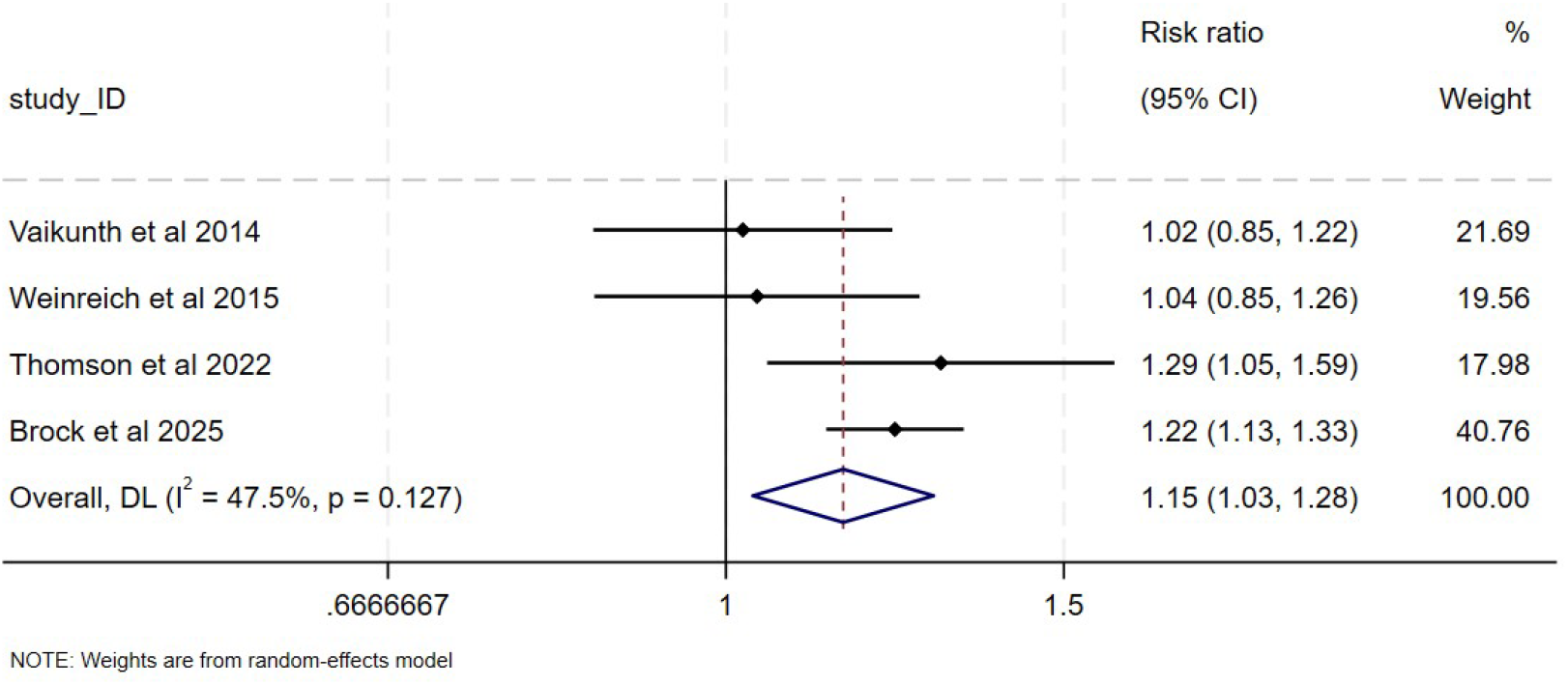

**Sensitivity Analysis 2:** Brown 2017 and Shah 2018 removed

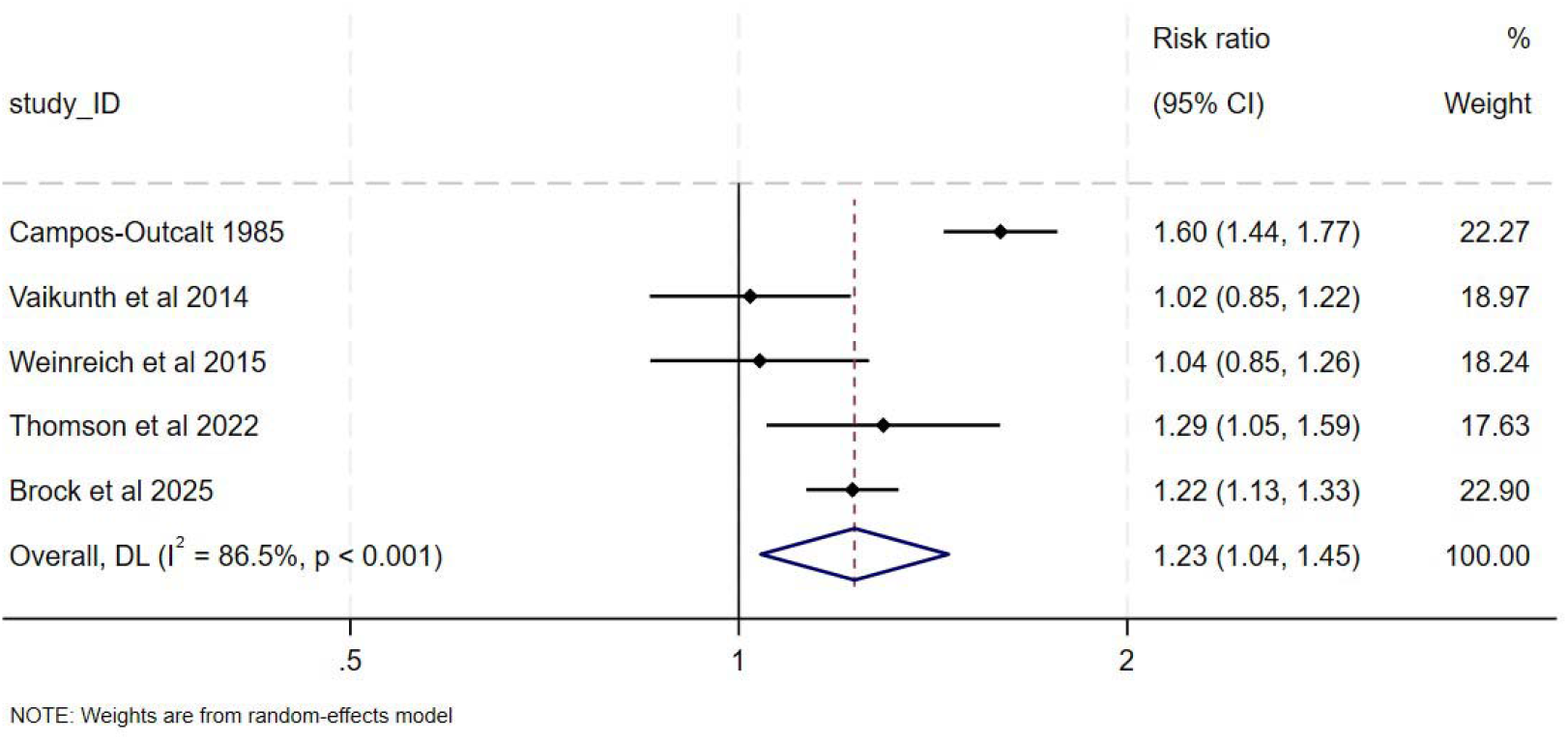

**Sensitivity Analysis 3:** Brown 2017 removed

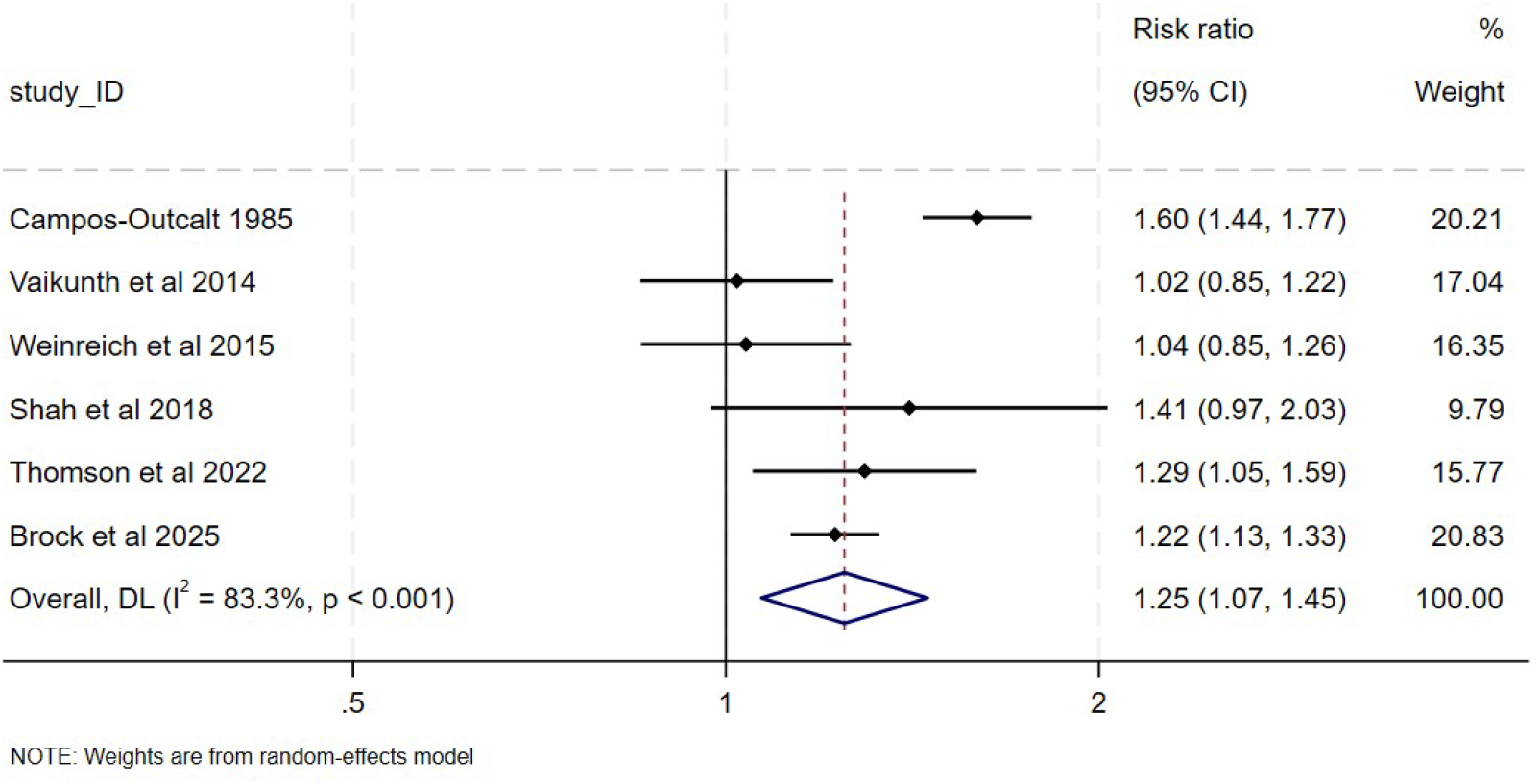

**Sensitivity Analysis 4:** Campos-Outcault 1985 removed

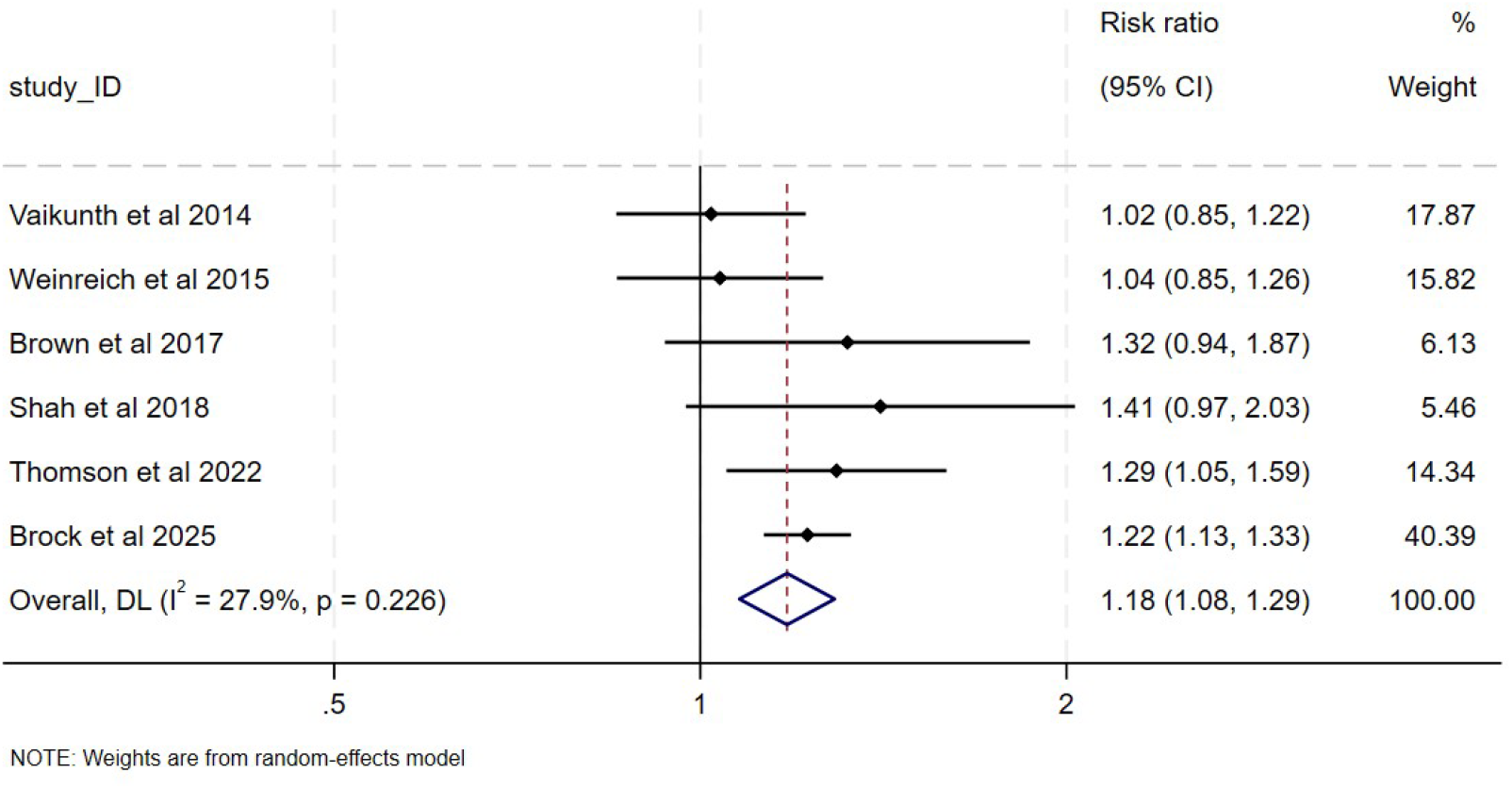

**Sensitivity Analysis 5:** Shah 2018 removed

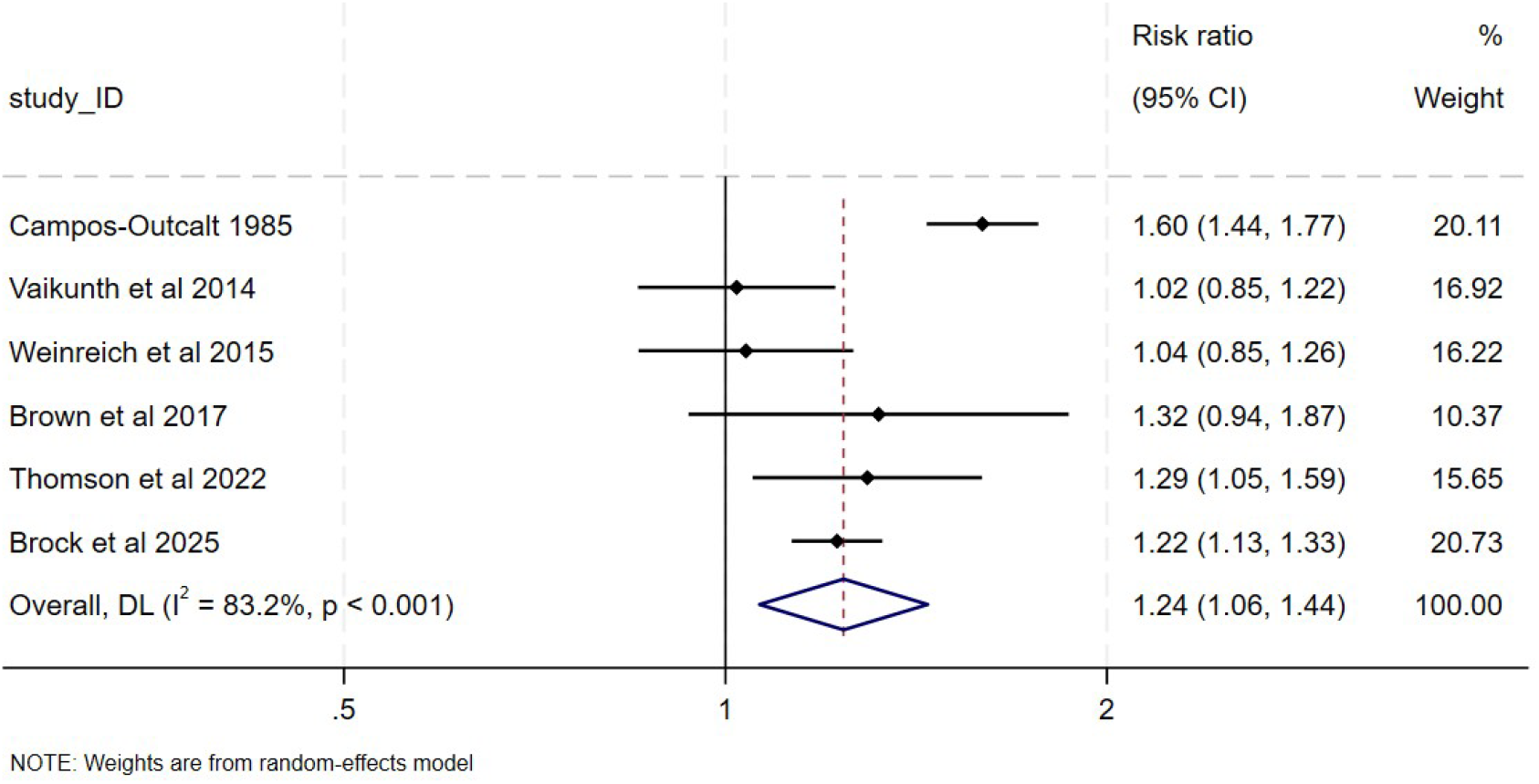

**Sensitivity Analysis 6:** Brock 2026 removed

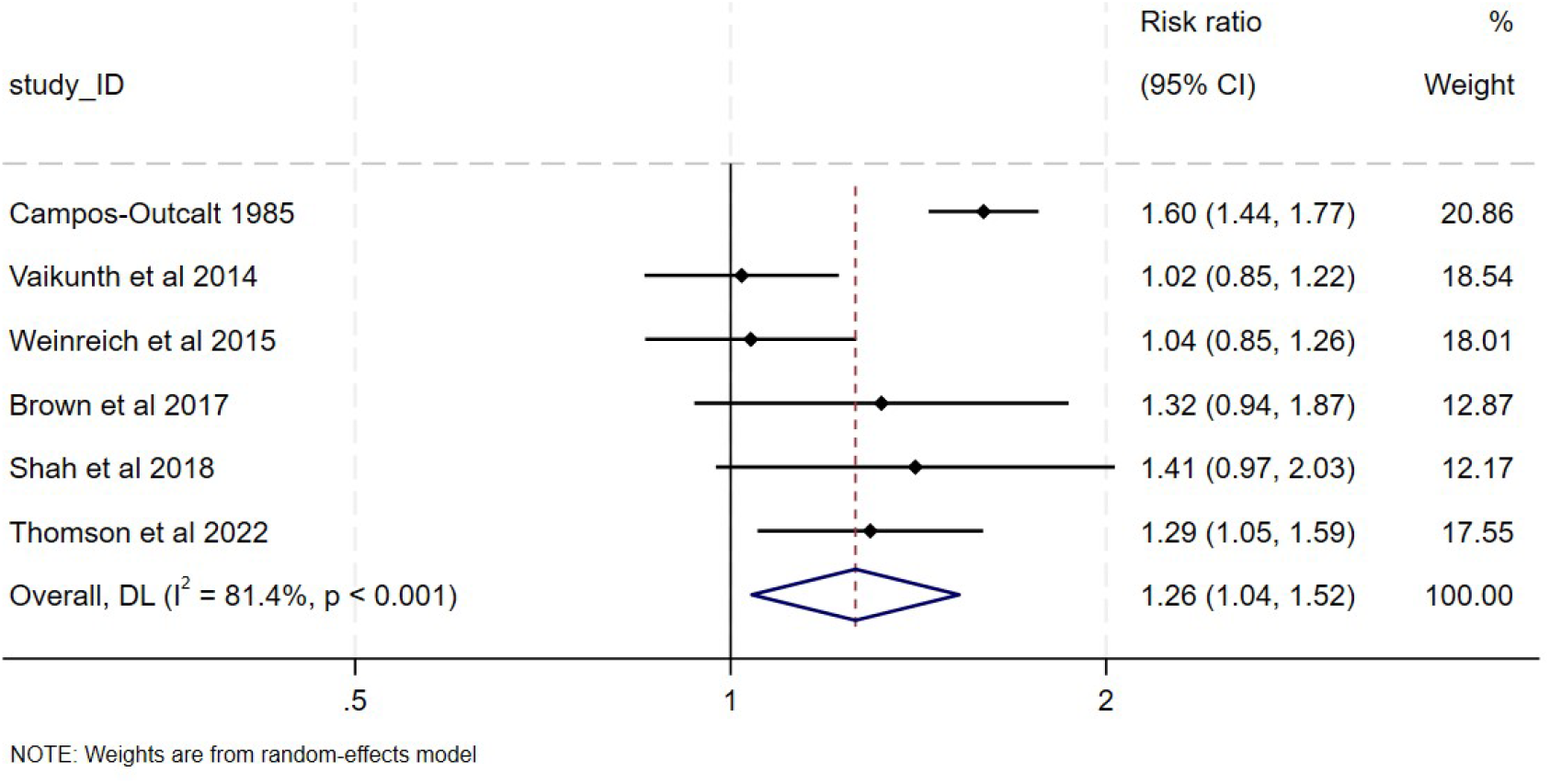

## Footnotes

a Inclusive of included and excluded studies on this topic

